# Guideline Adherence Following On-Site Versus Telestroke Consultation for Stroke Due to Intracranial Atherosclerosis

**DOI:** 10.64898/2026.08.06.26359874

**Authors:** Dawson Cooper, Abhiram Pillai, Elizabeth Harty, Natalia Crimmel, Samuel Worrell, Annette Xenopoulos-Oddsson, Erjia Cui, Praveen Hariharan, Margy McCullough-Hicks

**Affiliations:** Department of Neurology, Johns Hopkins School of Medicine, Baltimore, MD, USA; Department of Neurology, Cerebrovascular Division, University of Minnesota, Minneapolis, MN, USA; Masonic Institute for the Developing Brain, University of Minnesota, Minneapolis, MN, USA; Division of Biostatistics & Health Data Science, School of Public Health, University of Minnesota, Minneapolis, MN, USA

## Abstract

**Background:** Telestroke evaluation and treatment programs are a promising option for geographically underserved populations. Adherence to guideline-based secondary prevention measures among telestroke programs remains understudied, particularly for patients with symptomatic intracranial atherosclerosis. The primary objective of this study was to evaluate whether routine telestroke consultation provides guideline-concordant management comparable to on-site vascular neurology consultation.

**Methods:** This retrospective cohort review identified patients with stroke due to intracranial atherosclerosis within a single healthcare system comprising nine hospitals, including two comprehensive stroke centers with on-site stroke coverage and seven sites with remote telestroke coverage. Data was collected from January 2019 to December 2023. Adherence to guideline-based quality indicators was determined using four primary outcome measures including rates of permissive hypertension, high-intensity statin prescription at discharge, time to initiation of first antiplatelet medications, and appropriate antithrombotic therapy at discharge.

**Results:** A total of 132 patients were included in the final analysis (median age, 69 years; 65 female [49.2], 67 male [50.8%]), with 87 patients evaluated and managed on-site and 45 via telestroke. Guideline adherence was similar between groups for permissive hypertension and discharge antithrombotic therapy. Patients managed via telestroke were more likely to receive high-intensity statins at discharge (absolute difference 27.1% (95% CI 11.4, 42.8)) and received antiplatelet therapy earlier than patients managed on-site.

**Conclusion:** In this multisite, single-system cohort, routine telestroke consultation was associated with similar or greater adherence to selected guideline-based management measures compared with on-site vascular neurology consultation.

**KEY MESSAGES:** *What is already known on this topic:* Telestroke networks provide stroke expertise to rural areas.

*What this study adds:* Routine telestroke consultation for stroke due to intracranial atherosclerosis was associated with similar or greater adherence to selected guideline-based management measures compared with on-site vascular neurology consultation.

*How this study might affect research, practice, or policy:* Expanding routine telestroke coverage may reduce geographic disparities in access to high-quality, guideline-concordant vascular neurology care.

## INTRODUCTION

Stroke is a leading cause of morbidity and mortality worldwide^1^. Timely acute management, including intravenous thrombolysis and endovascular thrombectomy, improve functional outcomes considerably^2,3^. Access to these interventions and specialized medical management is largely limited to metropolitan areas with comprehensive stroke centers (CSCs), raising significant challenges for rural communities. Specific challenges for patients treated outside these centers include increased door to needle and door-in-door-out times resulting in higher rates of mortality^4–7^. Telemedicine-based stroke networks have emerged as a viable option for these communities, improving access to expert consultation and acute interventions.

Despite the growing use of these programs, studies regarding adherence to guideline-based best practices in the teleneurology setting for stroke remain limited. Initial data has been supportive of the integration of teleneurology-based services within the acute and subacute hospital setting^8^. Further evaluation, however, is needed to determine whether these findings are replicable among different stroke subtypes. Whether routine telestroke can deliver inpatient guideline-based secondary prevention comparable to in-person comprehensive stroke center vascular neurology services remains unknown.

Of the stroke subtypes, intracranial atherosclerosis (ICAS) carries the highest risk of early recurrence, providing a unique opportunity to study and optimize teleneurology-based stroke care^9–11^. Patients with stroke due to ICAS require implementation of several guideline-directed secondary prevention measures during hospitalization, making this population particularly well suited to evaluate adherence to evidence-based stroke care^12–15^. Determining whether teleneurology can reliably support the delivery of these therapies is essential as telestroke services continue to expand.

Here we compared adherence to guideline-based quality measures among patients with symptomatic ICAS managed by inpatient vascular neurologists at two CSCs versus the same vascular neurology team providing consultation through a telestroke network across seven affiliated community hospitals.

## METHODS

### Ethical Considerations and Data Sharing

This study was approved by the University of Minnesota Institutional Review Board (STUDY00023024). Aggregate data can be shared upon reasonable request to the principal investigator.

### Study Design and Patient Population

We performed an observational retrospective cohort review of patients hospitalized within the MHealth Fairview System at the University of Minnesota with stroke due to symptomatic ICAS between January 1st, 2019 and December 31st, 2023. MHealth Fairview is an integrated health system consisting of nine hospitals, including two academic tertiary referral centers for stroke care, one community hospital with inpatient, nonemergent stroke coverage, and six hospitals without any on-site stroke coverage. Further details regarding the setting and telestroke model have been published previously^16^. Vascular neurology consultation was provided by the same vascular neurology practice, including fellowship-trained vascular neurologists and advanced practice providers specializing in stroke, either through inpatient consultation at the comprehensive stroke centers or remotely through the telestroke networks.

This study included pre-specified inclusion and exclusion criteria that have been published previously^17^. Briefly, inclusion criteria included participants aged 18-89 and confirmed diagnosis of acutely symptomatic ICAS with 50-99% stenosis. This was confirmed by magnetic resonance angiography (MRA), computed tomography angiography (CTA), or cerebral angiogram along with the presence of an acute infarct in the corresponding territory on brain CT or MRI. Degree of stenosis was measured based on WASID (Warfarin-Aspirin Symptomatic Intracranial Disease criteria; 50%–69% versus 70%–99%) and location of culprit lesion (intracranial vertebral artery, basilar artery, intracranial internal carotid artery, M1 segment of the middle cerebral artery, or M2 segment of the middle cerebral artery)^18^. Exclusion criteria included acute endovascular treatment prior to a recurrent stroke, complete occlusion of the affected artery, hemorrhagic transformation with parenchymal hematoma, and presence of any acute infarct in a territory different from that of the affected artery.

### Primary Outcomes

Adherence to guideline-based quality indicators was determined using four primary outcome measures including permissive hypertension within the first 48 hours of hospitalization (no initiation of blood pressure medications within 48 hours from hospital arrival, except beta-blockers), high-intensity statin prescription (atorvastatin 80 mg or rosuvastatin 20 mg daily), time to initiation of first antiplatelet medication(s) (aspirin and clopidogrel), and appropriate antithrombotic therapy on discharge following stroke. Appropriate antithrombotic therapy was determined by 1) the degree of stenosis (50%–69% versus 70%–99%) and 2) the presence or absence of atrial fibrillation. For patients with 70-99% stenosis without atrial fibrillation, dual-antiplatelet therapy (DAPT) was appropriate. For patients with 50-69% stenosis without atrial fibrillation, single-antiplatelet therapy (SAPT) or DAPT was appropriate. For patients with atrial fibrillation, regardless of stenosis, SAPT alone was not appropriate. All other combinations were deemed as appropriate.

### Study Variables

Demographic variables included age at admission, biological sex, ethnicity, and patient reported race (White, Black, Asian, or other). Comorbid conditions included history of hypertension, history of diabetes, history of hyperlipidemia, history of atrial fibrillation, history of coronary artery disease, history of previous stroke, history of dementia, and current smoker status. Clinical variables included hours from last known well to arrival, systolic and diastolic blood pressure on admission, and national institutes of health score (NIHSS) on arrival. Imaging variables included degree of stenosis (50%–69% versus 70%–99%), imaging modality (MRA, CTA, or cerebral angiogram), affected artery (intracranial vertebral artery, basilar artery, intracranial internal carotid artery, M1 segment of the middle cerebral artery, or M2 segment of the middle cerebral artery, and acute infarct pattern on MRI (embolic, borderzone, perforator, or borderzone with embolic and/or perforator). These infarct patterns have been described previously^17^. Treatment variables included time (hours) to antiplatelet therapy from arrival, statin therapy, appropriate antithrombotic therapy, and permissive hypertension. Other variables included telestroke versus non-telestroke site, hospital at admission, and hospital at discharge.

### Data Collection & Imaging Training

Data was collected from patient electronic medical records. Patients were identified using our health system’s *Get With The Guidelines–Stroke* (GWTG-Stroke) quality improvement database, which captured all hospitalized stroke patients during the study period. Stroke etiology is assigned for each patient through standardized manual chart review by trained abstractors. We subsequently performed an independent review of all patients coded as having large artery atherosclerosis, cryptogenic stroke, or “other” stroke etiology to identify those meeting the predefined study criteria for symptomatic ICAS. First, authors (EH, NC, SW) screened eligible participants to confirm ICAS stroke etiology. Once ICAS-related stroke was confirmed, authors (DC, AP, PH, MMH) reviewed neuroimaging for each patient and determined the degree of stenosis of the affected vessel corresponding to the territory of infarct. To ensure consistent and accurate labeling of the acute infarct pattern, authors (DC, AP, PH, MMH) completed training as outlined in Yaghi et al, 2025^17^. Variables with unavailable data were treated as missing, and analyses were performed using only available-case data for each outcome.

### Data Analysis

Continuous variables were summarized as medians with interquartile ranges; categorical variables were summarized as frequencies with percentages. Unadjusted comparisons between patients managed on-site and those managed via telestroke were performed using independent-samples Student’s *t* tests for continuous variables and χ^2^ or Fisher’s exact tests for categorical variables.

Binomial logistic regression was used to evaluate associations between consultation model and dichotomous outcomes (coded as yes or no) including permissive hypertension during the first 48 hours of hospitalization, prescription of high-intensity statin therapy at discharge, and appropriate antithrombotic therapy at discharge. Results were reported as adjusted odds ratios (aOR) with 95% confidence intervals (CI). Multivariable logistic regression was used to evaluate differences in time from hospital arrival to first aspirin and clopidogrel administration. Results were reported as adjusted mean differences (β) with 95% CI. All adjusted regression models included consultation type as the primary exposure with the following prespecified covariates based on clinical relevance: age, self-reported race, admission NIHSS score, and history of hypertension, hyperlipidemia, diabetes mellitus, and prior stroke.

We conducted our analyses using R version 4.6.1 (The R Foundation) and Jamovi version 2.7.38 (The Jamovi Project) and considered 2-tailed *P* values less than 0.05 to be statistically significant. Differences in outcome means were calculated along with 95% CIs.

## RESULTS

### Baseline Characteristics

After screening eligible participants, a total of 132 patients from 9 total hospitals were included in the final analyses. The median age was 69 years old, 49.2% were women, and the median NIHSS score on arrival was 2. Further patient characteristics including race, ethnicity, comorbidities, and neuroimaging findings are reported in Table 1.

**Table 1.** Characteristics of study participants. Abbreviations: NIHSS, National Institutes of Health Stroke Scale; ICA, internal carotid artery; M1, M1 segment of the middle cerebral artery; M2, segment of the middle cerebral artery. *In some cases, the location of stenosis was noted in multiple arteries and reported as combination.

| Participant Characteristics | No. (%) |
| --- | --- |
| Patients, n | 132 |
| Demographics |  |
| Age, median (IQR) | 69 (19) |
| Sex |  |
| Female | 65 (49.2) |
| Male | 67 (50.8) |
| Ethnicity |  |
| Hispanic or Latino | 2 (1.5) |
| Non-Hispanic or Latino | 123 (93.2) |
| Not reported | 7 (5.3) |
| Race |  |
| White | 87 (65.9) |
| Black | 16 (12.1) |
| Asian | 22 (16.7) |
| Other | 5 (3.8) |
| Unknown | 2 (1.5) |
| NIHSS score, median (IQR) | 2 (3) |
| Comorbidities |  |
| Hypertension | 104 (78.8) |
| Diabetes | 59 (44.7) |
| Hyperlipidemia | 84 (63.7) |
| Coronary heart disease | 22 (16.8) |
| Congestive heart failure | 7 (5.3) |
| Atrial fibrillation | 5 (3.8) |
| History of stroke | 15 (11.4) |
| Active smoking | 22 (16.7) |
| History of dementia | 10 (7.6) |
| Degree of stenosis |  |
| 50-69% | 63 (47.7) |
| 70-99% | 69 (52.3) |
| Location of stenosis |  |
| ICA | 14 (10.6) |
| M1 | 45 (34.1) |
| M2 | 26 (19.7) |
| Vertebral | 14 (10.6) |
| Basilar | 28 (21.2) |
| Combination* | 5 (3.8) |
| Acute infarct pattern on MRI |  |
| Borderzone | 33 (25.6) |
| Embolic | 22 (17.1) |
| Perforator | 45 (34.9) |
| Borderzone with embolic and/or perforator | 29 (22.5) |

Demographic and other patient characteristics were then compared between patients evaluated and managed on-site versus telestroke. A total of 87 patients were evaluated and treated by on-site vascular neurologists. The remaining 45 patients were evaluated and treated remotely via telestroke by the same group of vascular neurologists throughout the study period. Overall, baseline characteristics were well balanced between groups, with no significant differences in demographics, vascular risk factors, stroke severity, or stenosis severity (Table 2). The only baseline difference between groups was acute infarct pattern (χ2 (3, N=129) = 8.28, p < 0.05), with combination infarct patterns occurring more frequently among patients managed on-site.

**Table 2.** Characteristics of study participants treated on-site versus those treated via telestroke services. Abbreviations: NIHSS, National Institutes of Health Stroke Scale; ICA, internal carotid artery; M1, M1 segment of the middle cerebral artery; M2, segment of the middle cerebral artery. *In some cases, the location of stenosis was noted in multiple arteries and reported as combination.

| Characteristics | On-Site, No. (%) | Telestroke, No. (%) | P value |
| --- | --- | --- | --- |
| Patients | 87 (65.9) | 45 (34.1) |  |
| Demographics |  |  |  |
| Age, median (IQR) | 68 (19) | 73 (19) | 0.422 |
| Female sex | 44 (50.6) | 21 (46.7) | 0.670 |
| Ethnicity: Hispanic or Latino | 1 (1.1) | 1 (2.2) | 0.601 |
| Race |  |  |  |
| White | 57 (65.5) | 30 (66.7) | 0.153 |
| Black | 12 (13.8) | 4 (8.9) |  |
| Asian | 11 (12.6) | 11 (24.4) |  |
| Other | 5 (5.7) | 0 (0.0) |  |
| NIHSS score, median (IQR) | 3 (3.0) | 1 (3.0) | 0.158 |
| Comorbidities, n (%) |  |  |  |
| Hypertension | 68 (78.2) | 36 (80.0) | 0.806 |
| Diabetes | 37 (42.5) | 22 (48.9) | 0.486 |
| Hyperlipidemia | 50 (57.5) | 34 (75.6) | 0.406 |
| Coronary heart disease | 16 (18.4) | 6 (13.3) | 0.443 |
| Congestive heart failure | 3 (3.4) | 4 (8.9) | 0.186 |
| Atrial fibrillation | 2 (2.3) | 3 (6.7) | 0.213 |
| History of stroke | 10 (11.5) | 5 (11.1) | 0.948 |
| Active smoking | 16 (18.4) | 6 (13.3) | 0.460 |
| History of dementia | 7 (8.0) | 3 (6.7) | 0.777 |
| Imaging Findings |  |  |  |
| Stenosis severity |  |  |  |
| 50-69% | 41 (47.1) | 22 (48.9) | 0.848 |
| 70-99% | 46 (52.9) | 23 (51.1) |  |
| Stenosis location |  |  |  |
| ICA | 7 (8.0) | 7 (15.5) | 0.565 |
| M1 | 32 (36.8) | 13 (28.9) |  |
| M2 | 19 (21.8) | 7 (15.5) |  |
| Vertebral | 8 (9.2) | 6 (13.3) |  |
| Basilar | 16 (18.4) | 12 (26.7) |  |
| *Combination | 5 (5.7) | 0 (0.0) |  |
| Acute infarct pattern on MRI |  |  |  |
| Borderzone | 20 (23.3) | 13 (30.2) | 0.041 |
| Embolic | 11 (12.8) | 11 (25.6) |  |
| Perforator | 30 (34.9) | 15 (34.9) |  |
| Borderzone with embolic and/or perforator | 25 (29.1) | 4 (9.3) |  |

### Adherence to Guideline-Based Measures

There were four primary guideline-based adherence measures included in this study: permissive hypertension during the first 48 hours of admission, high-intensity statin therapy at discharge, appropriate antithrombotic therapy at discharge, and time from arrival to first antiplatelet(s) administration.

There was no significant difference in rates of permissive hypertension adherence between on-site versus telestroke-managed patients (absolute difference 8.6%; 95% CI, -26.5 to 9.4) (Figure 1A). This finding was consistent when controlling for prespecified model covariates known to affect stroke management and outcome (aOR=0.72; 95% CI 0.323, 1.60) (Supplemental Table 1). Similarly, when comparing rates of appropriate antithrombotic therapy at discharge, no significant differences were found between on-site versus telestroke management (absolute difference 1.0%; 95% CI -8.13, 6.14) (Figure 1A); this finding was consistent when correcting for covariates (aOR=0.368; 95% CI 0.042, 3.22) (Supplemental Table 2).

**Figure 1.**
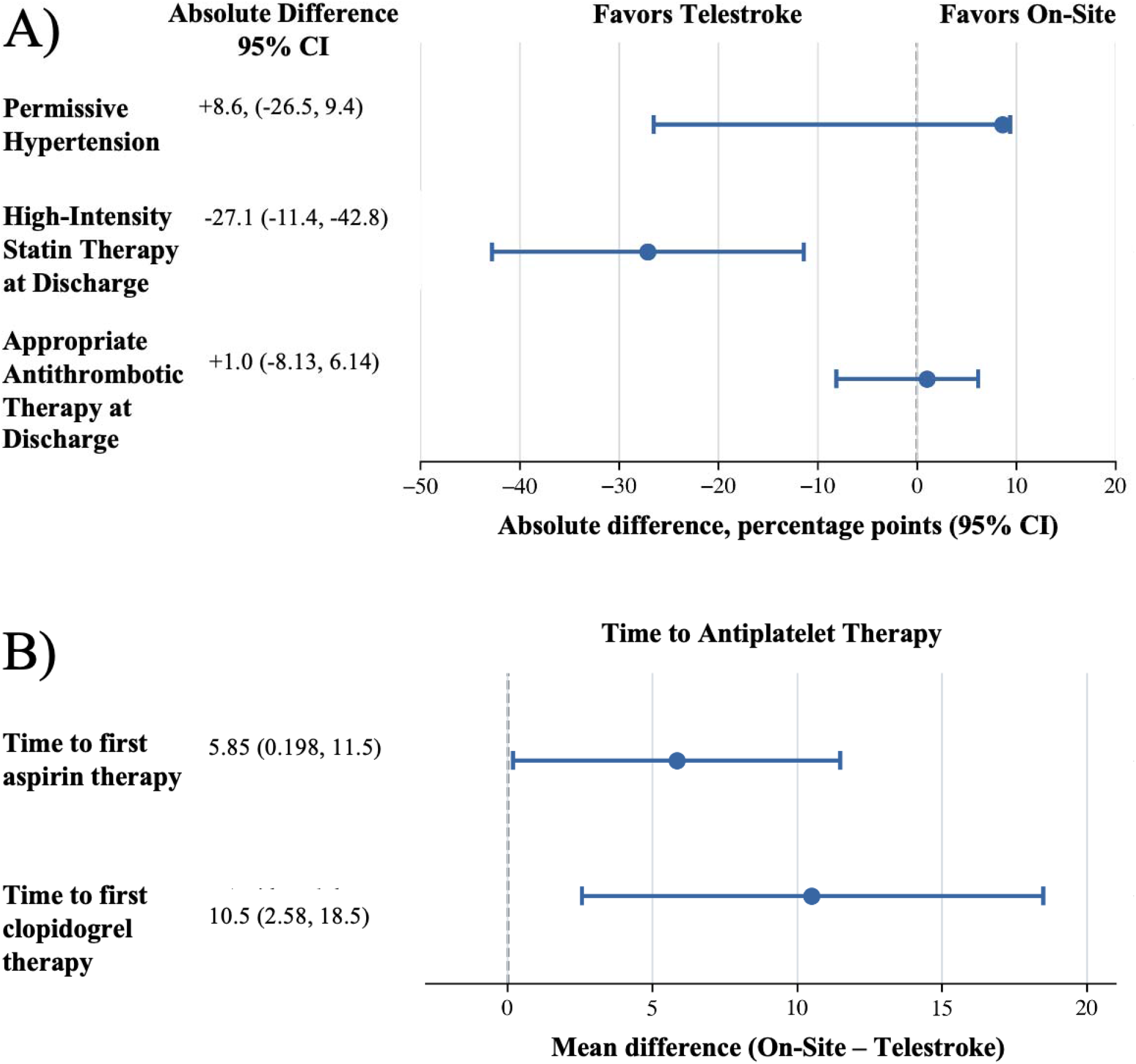
Guideline adherence in patients managed by inpatient vascular neurology versus telestroke consultation. Panel A) Absolute differences in adherence to guideline-based quality measures, expressed in percentage points. Panel B) Mean differences in time from hospital arrival to initiation of aspirin or clopidogrel therapy, expressed in hours. All estimates were calculated as on-site minus telestroke.

In contrast, when comparing on-site versus telestroke adherence for high-intensity statin therapy at discharge, there was a significant difference in favor of telestroke management. On-site adherence was 52.9% versus 80% of telestroke-managed cases (absolute difference 27.1%; 95% CI 11.4, 42.8) (Figure 1A). When correcting for covariates, telestroke remained independently associated with a higher odds of adherence compared to on-site management (aOR=4.437; 95% CI 1.765, 11.16) (Supplemental Table 3).

When comparing on-site versus telestroke management for time from arrival (hours) to first dose of aspirin, patients managed via telestroke were significantly more likely to receive their first dose of aspirin sooner compared to those treated on-site (absolute difference 5.85 hours; 95% CI 0.198, 11.5) (Figure 1B); however, when correcting for covariates, this finding did not remain statistically significant (β=-5.729; 95% CI -11.69, 0.232) (Supplemental Table 4). A similar analysis was conducted for time from arrival to first dose of clopidogrel, with telestroke-managed patients receiving their first dose of clopidogrel sooner compared to those treated on site (absolute difference 10.5 hours; 95% CI 2.58, 18.5) (Figure 1B). When correcting for covariates, this finding remained statistically significant (β=-11.38; 95% CI -19.87, -2.922) (Supplemental Table 5). An exploratory analysis excluding participants who received IV thrombolysis yielded similar findings for both time to aspirin and clopidogrel administration (Supplemental Tables 6 and 7, respectively).

## DISCUSSION

In this observational retrospective cohort review, we provide evidence that telestroke-based management of patients with stroke due to ICAS was comparable to, and for selected measures exceeded, on-site management across multiple guideline-based quality indicators. There were no significant differences between telestroke and on-site management for permissive hypertension within the first 48 hours of hospital admission as well as adherence to appropriate antithrombotic therapy on discharge. Adherence to high-intensity statin therapy was significantly higher among telestroke-managed patients. Telestroke-managed patients received their first dose of aspirin and clopidogrel significantly sooner than patients managed on-site. However, after adjustment for covariates known to affect stroke severity, time to first aspirin administration did not differ significantly between the groups. Overall, these findings support the continued integration of telestroke-based care for patients with stroke due to ICAS, especially in underserved areas with limited access to on-site vascular neurology expertise. Importantly, inpatient and telestroke consultations were provided by the same vascular neurology group, suggesting that adherence to guideline-based care can be maintained when expertise is delivered remotely rather than in person.

The goal of this study was to compare two real-world models of stroke care rather than isolate the effect of individual physicians or hospitals. Because the same vascular neurologists staffed both the inpatient vascular neurology service and the telestroke network, differences in adherence likely reflect the effectiveness of the overall care delivery model, including implementation of specialist recommendations within each clinical environment. These findings therefore support the ability of integrated telestroke systems to extend non-emergent comprehensive stroke expertise to hospitals without onsite vascular neurologists.

Prior research of single medical systems comparing telestroke versus standard on-site management has focused largely on the acute phase of stroke treatment, particularly door to needle times for intravenous thrombolysis^4,6,7,19,20^. The four quality measures reported in this study build on this work and span both acute and subacute phases of stroke management.

Among the reported guideline-based quality measures, acute management adherence was assessed using permissive hypertension within the first 48 hours of admission and time to first antiplatelet therapy. The 2025 AHA/ACC guideline recommends that for patients with acute ischemic stroke and BP <220/120 mm Hg who do not receive reperfusion therapy, active BP lowering within the first 48–72 hours is not effective to prevent death or dependency^13^. There were no significant differences in adherences between on-site and telestroke permissive hypertension management. For antiplatelet administration, telestroke-managed patients received clopidogrel more quickly than on-site managed participants. The same trend was found for aspirin administration; however, this finding was not statistically significant after covariate correction modeling. In practice, aspirin 160–300 mg within 24–48 hours is the standard antiplatelet therapy for acute ischemic stroke in patients not receiving thrombolysis; in those treated with IV thrombolysis, aspirin is delayed until 24 hours after administration due to the increased risk of hemorrhage following thrombolysis^15^. To assess potential confounding caused by thrombolysis administration, an exploratory analysis was conducted. This analysis excluded participants who received IV thrombolysis from both on-site and telestroke-managed patients. Results did not significantly differ between the analyses including and excluding those who received IV thrombolysis. Earlier antiplatelet administration may reflect the streamlined communication and standardized recommendations used within the telestroke consultation workflow, although this hypothesis warrants further study.

In the subacute phase, guideline-based quality measures included appropriate antithrombotic and high-intensity statin therapies on discharge. On-site and telestroke managed patients received appropriate antithrombotic therapy over 95% of the time, with no significant differences noted between the two groups. Adherence to high-intensity statin therapy, however, was significantly greater in telestroke-managed patients. The 2021 AHA/ASA guidelines strongly recommend high intensity stain therapy (atorvastatin 80 mg or rosuvastatin 20 mg daily) for secondary prevention following atherosclerotic ischemic stroke, with an LDL target <70 mg/dL^12,14^. Interestingly, a previous study found that teleneurology-based rounds were superior to on-site consultations with the most pronounced differences in secondary stroke prevention^8^. The authors concluded that this may, in part, reflect the frequent updates to guideline-based recommendations for secondary prevention. Because the same vascular neurologists provided care between treatment modalities, this is unlikely to contribute to our findings. One possible explanation is that telestroke consultations rely more on standardized recommendations and structured documentation, which may reduce practice variation and improve adherence to evidence-based secondary prevention measures. Additionally, on-site assessments may more frequently include providers in training with varying degrees of specialization. Nevertheless, our findings are generally in agreement with previous studies demonstrating support for telestroke management^8,21–23^.

### Strengths and Limitations

A major strength of this study is its focus on stroke due to symptomatic ICAS, which carries the highest risk of early recurrence among stroke etiologies^9–11^. To our knowledge, this is the first study to compare telestroke versus on-site management for a specific stroke subtype. The standardized guideline-based quality measures reported here are easily translatable to other stroke etiologies and can be replicated in future studies to better characterize telestroke management. The study also included guideline-based metrics encompassing multiple acuities (acute and subacute) of care within a single hospital stay for each participant, in contrast to focusing primarily on acute treatment. Another major strength is that both care models were staffed by the same vascular neurology group, minimizing provider-level variability and allowing direct comparison of the care delivery models themselves.

Several limitations should be considered when interpreting the results of this study. First, this was an observational retrospective study; the utility and caveats of this study type have been previously described^24^. Second, the relatively small sample size, particularly within the telestroke cohort, may have limited statistical power to detect differences in some guideline-based quality measures. Third, although previous studies have shown adherence to the quality measures reported here to improve patient outcomes, no such conclusion can be drawn regarding outcomes in our patient population as the study was not originally designed to assess clinical outcomes^12– 15^. Furthermore, quality of care cannot be entirely defined by the standardized indicators used here. The role of in-person communication and management among on-site team members should not be overlooked. Fourth, patients included in this study had a median NIHSS score of 2, limiting generalizability to more severe stroke presentations. Generalizability was also limited in this study as all participant data was collected from a single hospital system in the Midwest, which may not entirely reflect the patient populations of other systems and regions within the United States or internationally. Because this study was conducted within a single integrated health system using one vascular neurology group, results may not generalize to telestroke networks with different staffing models or clinical workflows.

## CONCLUSION

In conclusion, our study demonstrates that telestroke-managed patients with stroke due to ICAS received high-quality, guideline-adherent care comparable to inpatient vascular neurology consultation, with higher adherence observed for selected secondary prevention measures. These findings support the continued implementation of telestroke services, particularly to underserved regions with limited access to vascular neurology expertise. Future research should focus on including patients with more severe stroke symptoms and prospective evaluation of adherence to guideline-based quality indicators and their association with clinical outcomes (functional status, disability, mortality, quality of life). Furthermore, the cost-effectiveness of such programs compared to standard on-site care should be evaluated in the context of healthcare worker satisfaction.

## Data Availability

All data produced in the present study are available upon reasonable request to the authors.

## Data availability statement

Data are available upon reasonable request to corresponding author.

## Ethics Statements

### Patient consent for publication

Not applicable.

### Ethics approval

This study involves human participants. This study was approved by the University of Minnesota Institutional Review Board (STUDY00023024).

## Acknowledgements & Funding

All authors report no conflict of interest. Funding and support provided by the Masonic Institute for the Developing Brain and the Department of Neurology, University of Minnesota.

