## Supplemental Materials for "Guideline Adherence Following On-Site Versus Telestroke Consultation for Stroke Due to Intracranial Atherosclerosis"

| **Predictor** | **Adjusted Odds Ratio** | **95% Confidence Interval** | | **P value** |
| --- | --- | --- | --- | --- |
|  |  | **Upper** | **Lower** |  |
| Age | 0.988 | 0.957 | 1.02 | 0.450 |
| Site (0 = On-site, 1 = Telestroke) |  |  |  |  |
| 1 – 0 | 0.720 | 0.323 | 1.60 | 0.421 |
| Sex (0 = male, 1 = female): |  |  |  |  |
| 1 – 0 | 1.231 | 0.582 | 2.60 | 0.587 |
| Race (0 = White, 1 = Black, 2 = Asian, 3 = Other) |  |  |  |  |
| 1 – 0 | 1.737 | 0.492 | 6.14 | 0.391 |
| 2 – 0 | 1.065 | 0.345 | 3.28 | 0.913 |
| 3 – 0 | 2.571 | 0.234 | 28.24 | 0.440 |
| Hypertension (0 = No, 1 = Yes): |  |  |  |  |
| 1 – 0 | 1.655 | 0.610 | 4.49 | 0.323 |
| Diabetes (0 = No, 1 = Yes): |  |  |  |  |
| 1 – 0 | 0.429 | 0.177 | 1.04 | 0.061 |
| Hyperlipidemia (0 = No, 1 = Yes): |  |  |  |  |
| 1 – 0 | 1.748 | 0.692 | 4.42 | 0.238 |
| History of stroke (0 = No, 1 = Yes): |  |  |  |  |
| 1 – 0 | 1.128 | 0.339 | 3.76 | 0.845 |
| NIHSS score on hospital arrival | 0.984 | 0.880 | 1.10 | 0.776 |

**Supplemental Table 1.** Binomial logistic regressions results of adherence to permissive hypertension within the first 48 hours of hospitalization. Abbreviations: NIHSS, National Institutes of Health Stroke Scale.

| **Predictor** | **Adjusted Odds Ratio** | **95% Confidence Interval** | | **P value** |
| --- | --- | --- | --- | --- |
|  |  | **Upper** | **Lower** |  |
| Age | 1.037 | 0.953 | 1.13 | 0.401 |
| Site (0 = On-site, 1 = Telestroke) |  |  |  |  |
| 1 – 0 | 0.368 | 0.042 | 3.22 | 0.366 |
| Sex (0 = male, 1 = female): |  |  |  |  |
| 1 – 0 | 0.181 | 0.015 | 2.25 | 0.183 |
| Race (0 = White, 1 = Black, 2 = Asian, 3 = Other): |  |  |  |  |
| 1 – 0 | 4.58e+7 | 0.000 | Inf | 0.997 |
| 2 – 0 | 7.12e+7 | 0.000 | Inf | 0.996 |
| 3 – 0 | 0.457 | 0.008 | 23.68 | 0.697 |
| Hypertension (0 = No, 1 = Yes): |  |  |  |  |
| 1 – 0 | 1.120 | 0.069 | 18.06 | 0.936 |
| Diabetes (0 = No, 1 = Yes): |  |  |  |  |
| 1 – 0 | 0.845 | 0.090 | 7.90 | 0.883 |
| Hyperlipidemia (0 = No, 1 = Yes): |  |  |  |  |
| 1 – 0 | 0.879 | 0.077 | 10.05 | 0.918 |
| history of stroke (0 = No, 1 = Yes): |  |  |  |  |
| 1 – 0 | 0.392 | 0.029 | 5.34 | 0.482 |
| NIHSS score on hospital arrival | 0.892 | 0.695 | 1.14 | 0.368 |

**Supplemental Table 2.** Binomial logistic regressions results of adherence to appropriate antithrombotic therapy. Abbreviations: NIHSS, National Institutes of Health Stroke Scale

| **Predictor** | **Adjusted Odds Ratio** | **95% Confidence Interval** | | **P value** |
| --- | --- | --- | --- | --- |
|  |  | **Upper** | **Lower** |  |
| Age | 0.973 | 0.939 | 1.01 | 0.121 |
| Site (0 = On-site, 1 = Telestroke) |  |  |  |  |
| 1 – 0 | 4.437 | 1.765 | 11.16 | 0.002 |
| Sex (0 = male, 1 = female): |  |  |  |  |
| 1 – 0 | 1.998 | 0.894 | 4.46 | 0.092 |
| Race (0 = White, 1 = Black, 2 = Asian, 3 = Other): |  |  |  |  |
| 1 – 0 | 3.538 | 0.794 | 15.76 | 0.097 |
| 2 – 0 | 1.178 | 0.339 | 4.10 | 0.797 |
| 3 – 0 | 4.022 | 0.361 | 44.84 | 0.258 |
| Hypertension (0 = No, 1 = Yes): |  |  |  |  |
| 1 – 0 | 1.890 | 0.655 | 5.45 | 0.239 |
| Diabetes (0 = No, 1 = Yes): |  |  |  |  |
| 1 – 0 | 0.639 | 0.244 | 1.67 | 0.360 |
| Hyperlipidemia (0 = No, 1 = Yes): |  |  |  |  |
| 1 – 0 | 1.652 | 0.651 | 4.20 | 0.291 |
| history of stroke (0 = No, 1 = Yes): |  |  |  |  |
| 1 – 0 | 0.545 | 0.160 | 1.86 | 0.333 |
| NIHSS score on hospital arrival | 0.977 | 0.869 | 1.10 | 0.702 |

**Supplemental Table 3.** Binomial logistic regressions results of adherence to high-intensity statin prescription on discharge. Abbreviations: NIHSS, National Institutes of Health Stroke Scale.

| **Predictor** | **Difference (β)** | **95% Confidence Interval** | | **P value** |
| --- | --- | --- | --- | --- |
|  |  | **Upper** | **Lower** |  |
| Age | 0.086 | -0.149 | 0.321 | 0.469 |
| Site (0 = On-site, 1 = Telestroke) |  |  |  |  |
| 1 – 0 | -5.729 | -11.69 | 0.232 | 0.060 |
| Sex (0 = male, 1 = female): |  |  |  |  |
| 1 – 0 | 0.523 | -4.962 | 6.008 | 0.851 |
| Race (0 = White, 1 = Black, 2 = Asian, 3 = Other): |  |  |  |  |
| 1 – 0 | 7.534 | -1.316 | 16.38 | 0.095 |
| 2 – 0 | 0.964 | -7.427 | 9.355 | 0.820 |
| 3 – 0 | -8.724 | -23.83 | 6.379 | 0.255 |
| Hypertension (0 = No, 1 = Yes): |  |  |  |  |
| 1 – 0 | -3.023 | -10.41 | 4.358 | 0.419 |
| Diabetes (0 = No, 1 = Yes): |  |  |  |  |
| 1 – 0 | -1.046 | -7.466 | 5.375 | 0.748 |
| Hyperlipidemia (0 = No, 1 = Yes): |  |  |  |  |
| 1 – 0 | 0.604 | -5.921 | 7.129 | 0.855 |
| history of stroke (0 = No, 1 = Yes): |  |  |  |  |
| 1 – 0 | 1.665 | -7.193 | 10.52 | 0.710 |
| NIHSS score on hospital arrival | 0.790 | -0.015 | 1.594 | 0.054 |

**Supplemental Table 4.** Multivariable logistic regression results of time from hospital arrival to first aspirin administration. Abbreviations: NIHSS, National Institutes of Health Stroke Scale.

| **Predictor** | **Difference (β)** | **95% Confidence Interval** | | **P value** |
| --- | --- | --- | --- | --- |
|  |  | **Upper** | **Lower** |  |
| Age in years | 0.288 | -0.047 | 0.622 | 0.091 |
| Site (0 = On-site, 1 = Telestroke) |  |  |  |  |
| 1 – 0 | -11.38 | -19.87 | -2.922 | 0.009 |
| Sex (0 = male, 1 = female): |  |  |  |  |
| 1 – 0 | -4.873 | -12.63 | 2.883 | 0.216 |
| Race (0 = White, 1 = Black, 2 = Asian, 3 = Other): |  |  |  |  |
| 1 – 0 | 1.267 | -10.75 | 13.29 | 0.836 |
| 2 – 0 | 1.180 | -10.41 | 12.78 | 0.841 |
| 3 – 0 | -13.94 | -35.58 | 7.692 | 0.205 |
| Hypertension (0 = No, 1 = Yes): |  |  |  |  |
| 1 – 0 | -0.672 | -10.78 | 9.435 | 0.895 |
| Diabetes (0 = No, 1 = Yes): |  |  |  |  |
| 1 – 0 | 6.843 | -2.319 | 16.00 | 0.142 |
| Hyperlipidemia (0 = No, 1 = Yes): |  |  |  |  |
| 1 – 0 | -2.467 | -11.56 | 6.625 | 0.592 |
| history of stroke (0 = No, 1 = Yes): |  |  |  |  |
| 1 – 0 | 6.262 | -7.412 | 19.94 | 0.366 |
| NIHSS score on hospital arrival | 0.639 | -0.556 | 1.833 | 0.291 |

**Supplemental Table 5.** Multivariable logistic regression results of time from hospital arrival to first clopidogrel administration. Abbreviations: NIHSS, National Institutes of Health Stroke Scale.

| **Predictor** | **Difference (β)** | **95% Confidence Interval** | | **P value** |
| --- | --- | --- | --- | --- |
|  |  | **Upper** | **Lower** |  |
| Age in years | 0.198 | -0.054 | 0.449 | 0.122 |
| Site (0 = On-site, 1 = Telestroke) |  |  |  |  |
| 1 – 0 | -2.617 | -8.459 | 3.224 | 0.376 |
| Sex (0 = male, 1 = female): |  |  |  |  |
| 1 – 0 | 0.201 | -5.373 | 5.776 | 0.943 |
| Race (0 = White, 1 = Black, 2 = Asian, 3 = Other): |  |  |  |  |
| 1 – 0 | 11.73 | 3.070 | 20.40 | 0.008 |
| 2 – 0 | 3.865 | -4.479 | 12.21 | 0.360 |
| 3 – 0 | -3.885 | -18.45 | 10.68 | 0.597 |
| Hypertension (0 = No, 1 = Yes): |  |  |  |  |
| 1 – 0 | -3.348 | -10.70 | 4.003 | 0.368 |
| Diabetes (0 = No, 1 = Yes): |  |  |  |  |
| 1 – 0 | -1.279 | -7.693 | 5.135 | 0.693 |
| Hyperlipidemia (0 = No, 1 = Yes): |  |  |  |  |
| 1 – 0 | -1.189 | -7.859 | 5.480 | 0.724 |
| history of stroke (0 = No, 1 = Yes): |  |  |  |  |
| 1 – 0 | 3.465 | -5.409 | 12.34 | 0.440 |
| NIHSS score on hospital arrival | 0.726 | -0.073 | 1.525 | 0.074 |

**Supplemental Table 6.** Multivariable logistic regression results of time from hospital arrival to first aspirin administration. All patients (n=12) who received thrombolytic treatment were excluded from the analysis. Abbreviations: NIHSS, National Institutes of Health Stroke Scale.

| **Predictor** | **Difference (β)** | **95% Confidence Interval** | | **P value** |
| --- | --- | --- | --- | --- |
|  |  | **Upper** | **Lower** |  |
| Age in years | 0.392 | 0.022 | 0.761 | 0.038 |
| Site (0 = On-site, 1 = Telestroke) |  |  |  |  |
| 1 – 0 | -9.348 | -18.08 | -0.619 | 0.036 |
| Sex (0 = male, 1 = female): |  |  |  |  |
| 1 – 0 | -5.381 | -13.66 | 2.898 | 0.200 |
| Race (0 = White, 1 = Black, 2 = Asian, 3 = Other): |  |  |  |  |
| 1 – 0 | 4.824 | -7.668 | 17.32 | 0.445 |
| 2 – 0 | 4.186 | -8.054 | 16.43 | 0.499 |
| 3 – 0 | -10.74 | -32.65 | 11.17 | 0.3329 |
| Hypertension (0 = No, 1 = Yes): |  |  |  |  |
| 1 – 0 | -0.954 | -11.57 | 9.660 | 0.859 |
| Diabetes (0 = No, 1 = Yes): |  |  |  |  |
| 1 – 0 | 7.170 | -2.392 | 16.73 | 0.140 |
| Hyperlipidemia (0 = No, 1 = Yes): |  |  |  |  |
| 1 – 0 | -3.980 | -13.64 | 5.689 | 0.416 |
| history of stroke (0 = No, 1 = Yes): |  |  |  |  |
| 1 – 0 | 8.851 | -5.118 | 22.82 | 0.212 |
| NIHSS score on hospital arrival | 0.443 | -0.833 | 1.718 | 0.493 |

**Supplemental Table 7.** Multivariable logistic regression results of time from hospital arrival to first clopidogrel administration. All patients (n=12) who received thrombolytic treatment were excluded from the analysis. Abbreviations: NIHSS, National Institutes of Health Stroke Scale.
